# The Modern Utility of Awake Deep Brain Stimulation Surgery

**DOI:** 10.1101/2025.09.25.25336654

**Authors:** Bryan T. Klassen, Matthew R. Baker, Kai J. Miller

**Author notes:** Correspondence: Bryan T. Klassen, MD, Department of Neurology, Mayo Clinic, 200 1^st^ ST SW, Rochester, MN, 55905. Bryan T. Klassen and Matthew R. Baker contributed equally to this work.

## Abstract

**Background and Objectives:** Advances in neuroimaging and stereotactic accuracy have led many centers to adopt asleep workflows for deep brain stimulation (DBS), raising questions about the continued role of awake surgery with microelectrode recording (MER). We evaluated whether intraoperative physiologic testing during awake ventral intermediate nucleus (Vim) DBS results in trajectory adjustments that converge with the imaging-defined target.

**Methods:** We retrospectively reviewed 137 consecutive Vim DBS leads implanted in awake patients. For each lead, we determined whether the final implant followed the planned trajectory, required a depth adjustment, or was moved to a parallel trajectory based on MER and test stimulation. In cases involving parallel trajectory adjustments, co-registered preoperative imaging and postoperative CT were used to compare the final lead position with the imaging-defined target and determine the direction and magnitude of deviation. Intraoperative ring stimulation data were analyzed to compare tremor suppression and side effects before and after trajectory adjustment.

**Results:** Of 137 leads, 116 (85%) were implanted along the planned trajectory, including 49 at the planned depth and 67 with depth adjustments. Twenty-one leads (15%) were moved to a parallel trajectory based on intraoperative findings; 17 had data suitable for spatial analysis. Among these, 13 (76%) were moved farther from the imaging-defined target, 2 (12%) an equal distance across the target, and 2 (12%) closer. In 12 cases with paired pre- and post-adjustment stimulation testing, trajectory adjustment was associated with reduced stimulation required for near tremor resolution, increased side-effect thresholds, and expansion of the therapeutic window. No leads required revision.

**Conclusion:** Intraoperative physiologic testing during awake Vim DBS frequently prompted trajectory adjustments that would not have been made under imaging guidance alone and were associated with improved stimulation response, underscoring the continuing role of awake testing in refining lead placement to patient-specific functional anatomy.

## INTRODUCTION

At inception, deep brain stimulation (DBS) for the treatment of movement disorders was reliant upon detailed atlases of inferred anatomic boundaries that could be understood when compared with anatomic landmarks identified on intraoperative ventriculography^1,2^. Intraoperative teams of neurosurgeons, neurologists and neurophysiologists conducted sophisticated multitrack studies of baseline cell firing patterns, changes in cell activity (firing rate) during sensory/motor tasks, and behavioral responses to test stimulation to infer the correct position for permanent deep brain stimulation electrode placement^3–5^. Generations of patients have benefited as a result.

In the past 35 years, dramatic breakthroughs in brain imaging including new sequences and multimodal fusion, stereotactic targeting accuracy, and the ability to obtain volumetric imaging before, during, and after DBS electrode placement have improved precision of electrode delivery to intended anatomic structures^6–8^. As a result, many centers have adopted asleep DBS workflows prioritizing imaging-defined endpoints and minimizing or eliminating intraoperative physiologic testing.

Despite these advances, uncertainty remains regarding how imaging-defined targets relate to patient-specific functional anatomy at the individual level. Imaging can identify candidate targets and surrounding structures but does not directly assess neuronal activity or stimulation-induced clinical effects. As a result, intraoperative physiologic findings and test stimulation may prompt adjustments that are not predicted by preoperative imaging alone. While randomized trials and meta-analyses often report similar short-term clinical outcomes between awake and asleep DBS, these studies are not designed to interrogate intraoperative decision-making or the factors that influence final lead placement.

The present study focuses on a fundamental question: when intraoperative physiologic testing during awake DBS prompts a change in lead placement, do adjustments move the lead closer to the imaging-defined target (correcting for stereotactic error), or do they diverge from it? By examining the direction and magnitude of physiologically guided adjustments, we aim to clarify how imaging-based planning and real-time physiologic feedback interact during contemporary DBS surgery. We address this question using ventral intermediate nucleus (Vim) DBS for essential tremor as an illustrative test context. Vim targeting integrates indirect stereotactic coordinates, atlas-based segmentation, tractography of the dentatorubrothalamic tract, and, in many practices, intraoperative physiologic confirmation.

We performed a retrospective review of consecutive awake Vim DBS cases performed by a single surgeon working with a single intraoperative neurologist. By categorizing final lead placement relative to the planned imaging-defined target, we quantified how often intraoperative physiology influenced trajectory or depth selection and to characterize the direction of those adjustments when they occurred.

## METHODS

To evaluate how intraoperative physiologic testing influences lead placement during awake DBS, we retrospectively reviewed consecutive ventral intermediate nucleus (Vim) DBS cases performed over a five-year interval by a single surgeon working with a single intraoperative neurologist. Vim DBS for essential tremor was selected to constrain analyses to a single anatomic target and clinical indication.

All procedures followed a standardized workflow that evolved incrementally over time with the integration of improved MRI sequences and atlas segmentations, but without substantive changes to the intraoperative decision-making framework. All our patients received a rechargeable system with segmented leads having a 1-3-3-1 pattern (levels are 1.5mm long, with 0.5mm between contact levels; Boston Scientific Corporation).

### Ethics statement

The study was conducted according to the Institutional Review Board of Mayo Clinic Rochester (IRB# 19-009878). Each patient provided informed written consent as approved by the IRB.

### Surgical planning

Preoperative selection of Vim thalamic targets was performed using Stealth (Medtronic, Inc.) or BrainLab (Brainlab AG) navigation software. High-resolution (voxel size ≤ 1 x 1 x 1 mm) 3T MR sequences including T1-weighted, T2-weighted, white-matter nulled (FGATIR), and DTI sequences were coregistered for multimodal planning. Our indirect target was on the AC-PC axis at a point posterior to mid-commissural point by 25% of the AC-PC distance and ∼10mm lateral to the wall of the 3^rd^ ventricle^9,10^. Each patient’s MRI (T1w, T2w sequences) was normalized into MNI space using the SyN registration approach as implemented in Advanced Normalization Tools as a five-stage nonlinear deformation^11^. MNI-space atlases depicting the Vim target^12^ and an ET tremor efficacy hotspot^13^ were intensity encoded on the patient’s preoperative MRI by applying the normalization matrix in reverse. This patient-specific atlas was co-registered with other pre-operative images in the planning software. Using deterministic tractography, tracts were calculated for the target circuit (DRTT seeded from the ipsilateral cerebellar nuclei and precentral gyrus) and for nearby circuits mediating side-effect (medial lemniscus seeded from the ipsilateral posterior midbrain and postcentral gyrus; corticospinal tract seeded from the ipsilateral precentral gyrus and cerebral peduncle)^14^. In many cases, the DRTT was directly visible on the white-matter nulled sequences (Fig. 1A).

**Figure 1.**
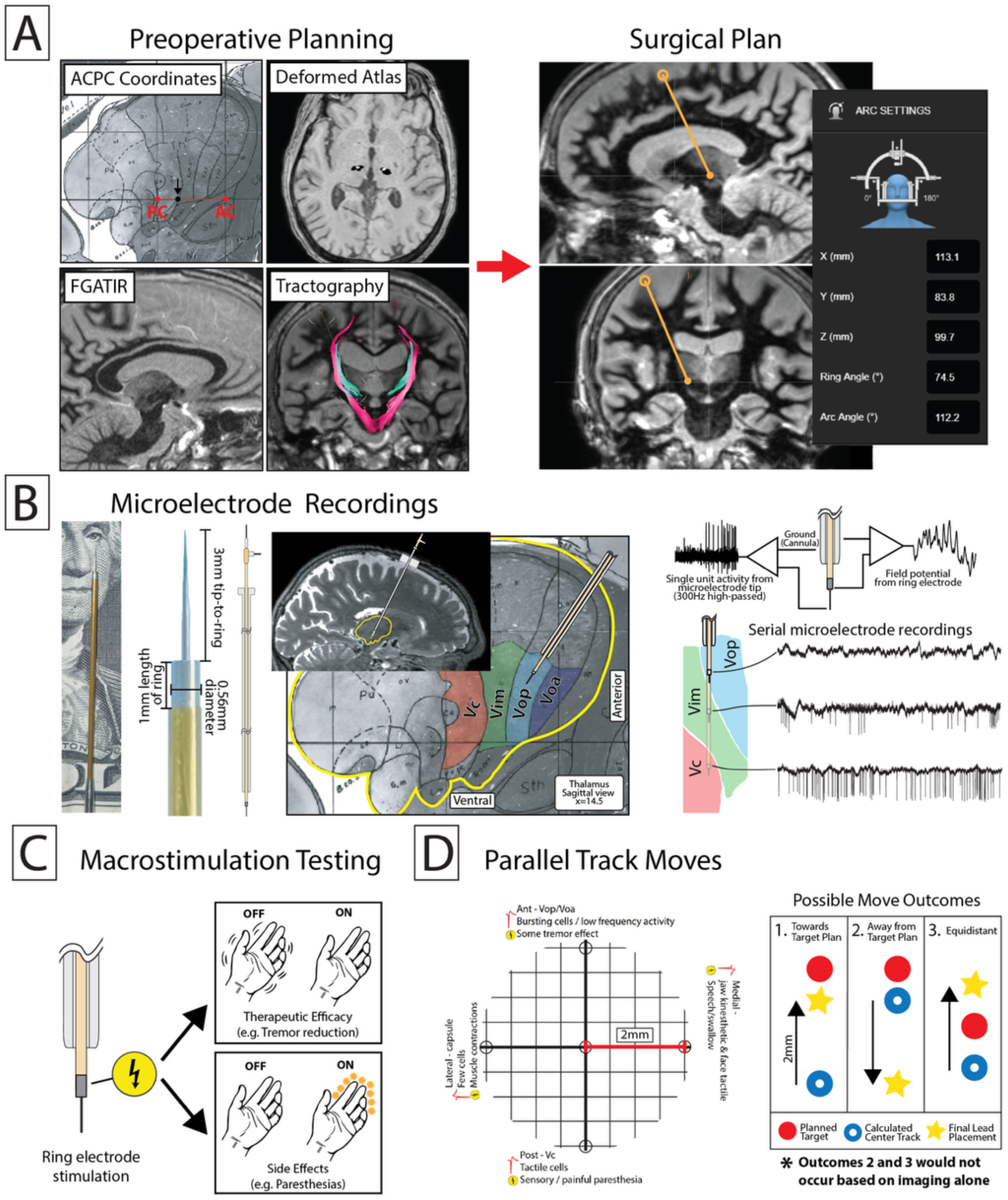
Awake deep brain stimulation (DBS) surgical workflow. **(A)** Preoperative planning integrates indirect targeting (ACPC coordinates), deformed atlases, and structural and diffusion imaging (FGATIR, tractography) to define surgical targets and trajectories. **(B)** Microelectrode recordings (MER) involves inserting high-impedance electrodes into target structures to record single-cell activity and local field potentials. **(C)** Macrostimulation testing uses the ring electrode to assess therapeutic efficacy and side effect thresholds to confirm or reposition the lead. **(D)** In suboptimal cases, additional tracks can be tested. These moves may be closer to the target, correcting for stereotactic error, or farther/equidistant moves that would typically not be made by imaging alone.

To select the final surgical target, we integrated the four above-described modalities. In cases where these did not converge on a single target, we gave more weight to modalities with higher quality and higher patient specificity (a well delineated DRTT on white-matter nulled imaging having the greatest weight, and AC-PC indirect targeting the least weight).

### Stereotactic implantation

Regularly calibrated Leksell G-frames (Elekta AB) were used for each surgery. The frame base was placed using local anesthesia in the preoperative area, and a mixed-phase contrast thin-cut zero-gantry volumetric CT was used for registration. This was coregistered to preoperative MRIs and the predetermined targets using the navigation software. Nearly all patients underwent bilateral implantation.

### Microelectrode recording and test stimulation

MER was performed using the NeuroOmega system (Alpha Omega Engineering). In cases where preoperative imaging modalities converged on a clear target, initial recordings were obtained along a single trajectory. When imaging inputs diverged or physiologic findings were ambiguous, recordings were obtained along multiple simultaneous trajectories.

A microelectrode was advanced through a rigid cannula in submillimeter increments toward target (Fig. 1B). Background activity, single-unit firing patterns, and task-related modulation were assessed to identify structural boundaries and functional correlates. Typically, measurements are recorded in 1mm increments for 30 seconds per site, resulting in approximately 15 sites and <10 minutes of recording time per track. Test stimulation was performed through the ring electrode on the cannula (3mm above the microelectrode tip), typically positioned 2 mm above the planned target to observe tremor improvement and side effects (Fig. 1C). If results of the observed microelectrode and test stimulation were as expected from the plan, the permanently implantable DBS lead was placed.

Next, bipolar stimulation was applied using the Boston Scientific external neurostimulator using a frequency of 130 Hz, pulse width of 60 microseconds, and step-wise increase in amplitude up to 3-4 mA. Patients were again evaluated for tremor improvement and stimulation-induced side effects. If the intraoperative neurologist thought there was not a sufficient therapeutic window between tremor resolution and side effect, an additional pass was made along a parallel trajectory 2 mm away (Fig. 1D), while keeping the original cannula in place to minimize displacement of brain tissue.

Figure 2 shows example MER traces including tremor- and passive movement-tuned cells (kinesthetic cells), which may indicate the Vim region, while sensory-tuned cells indicate sensory thalamus to be avoided.

**Figure 2.**
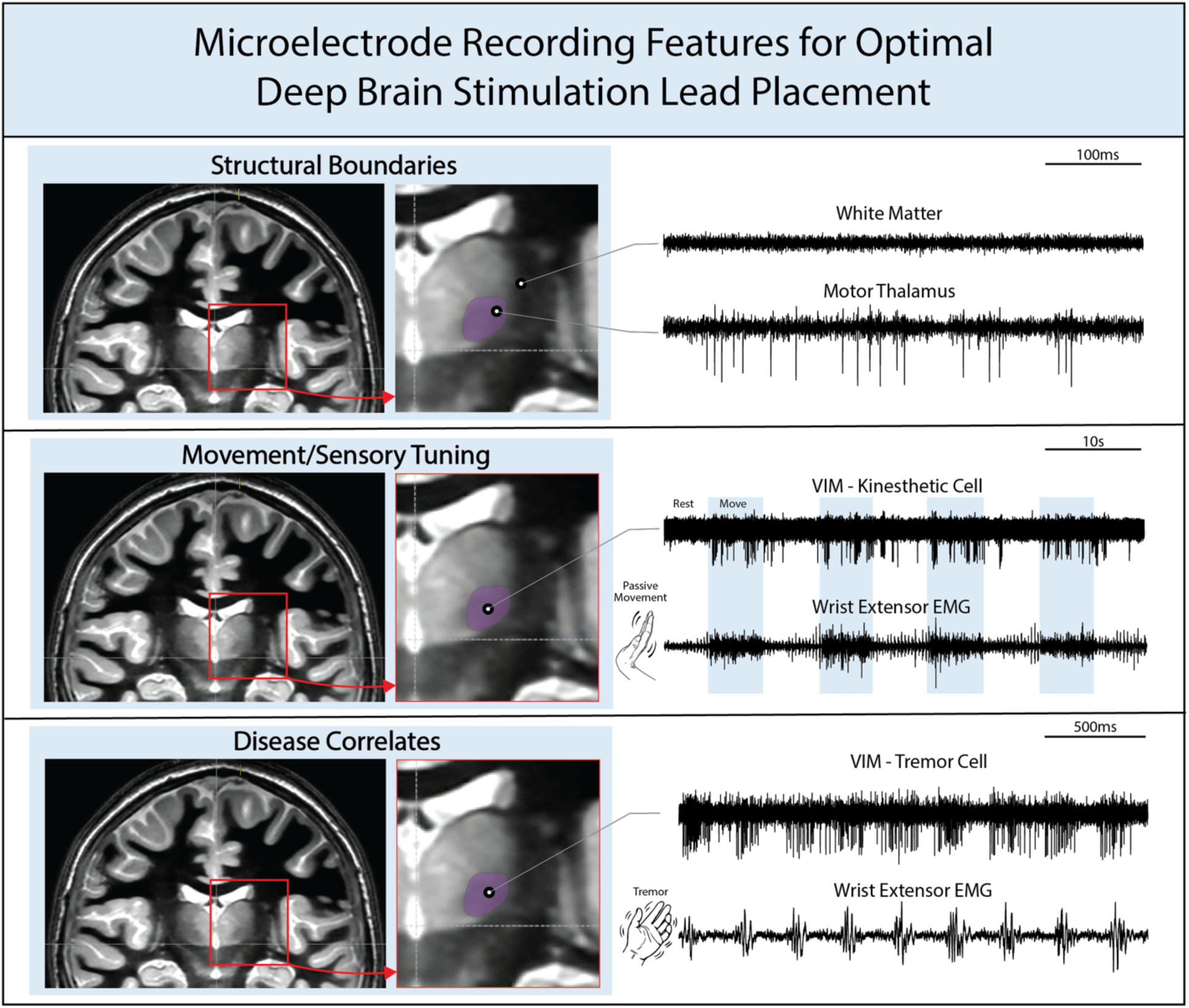
Features of microelectrode recordings (MER) used for optimal deep brain stimulation (DBS) lead placement in the ventral intermediate nucleus of the thalamus (Vim). MER identifies white to gray matter transitions based on single-unit firing and background (e.g. quiet white matter into highly active motor thalamus back into quiet white matter past ventral border). Neurons in the Vim may be tuned to passive movement (kinesthetic cells) or pathologic tremor as seen on electromyography (EMG).

### Postoperative imaging and lead localization

Postoperative head CT imaging was obtained on the day of surgery following the lead implantation procedure. For cases involving parallel trajectory adjustments, preoperative imaging was co-registered to postoperative CT, and the final lead position was determined based on the lead artifact (Fig. 3A-B). The position of the originally explored trajectory—also referred to as the calculated center trajectory—was reconstructed based on the known stereotactic offsets from the final lead position. Both final lead position and calculated center trajectory were compared with the image-defined plan to determine whether physiologically guided adjustments moved the lead closer to, farther from, or equidistant across the imaging-defined target (Fig. 3C).

**Figure 3.**
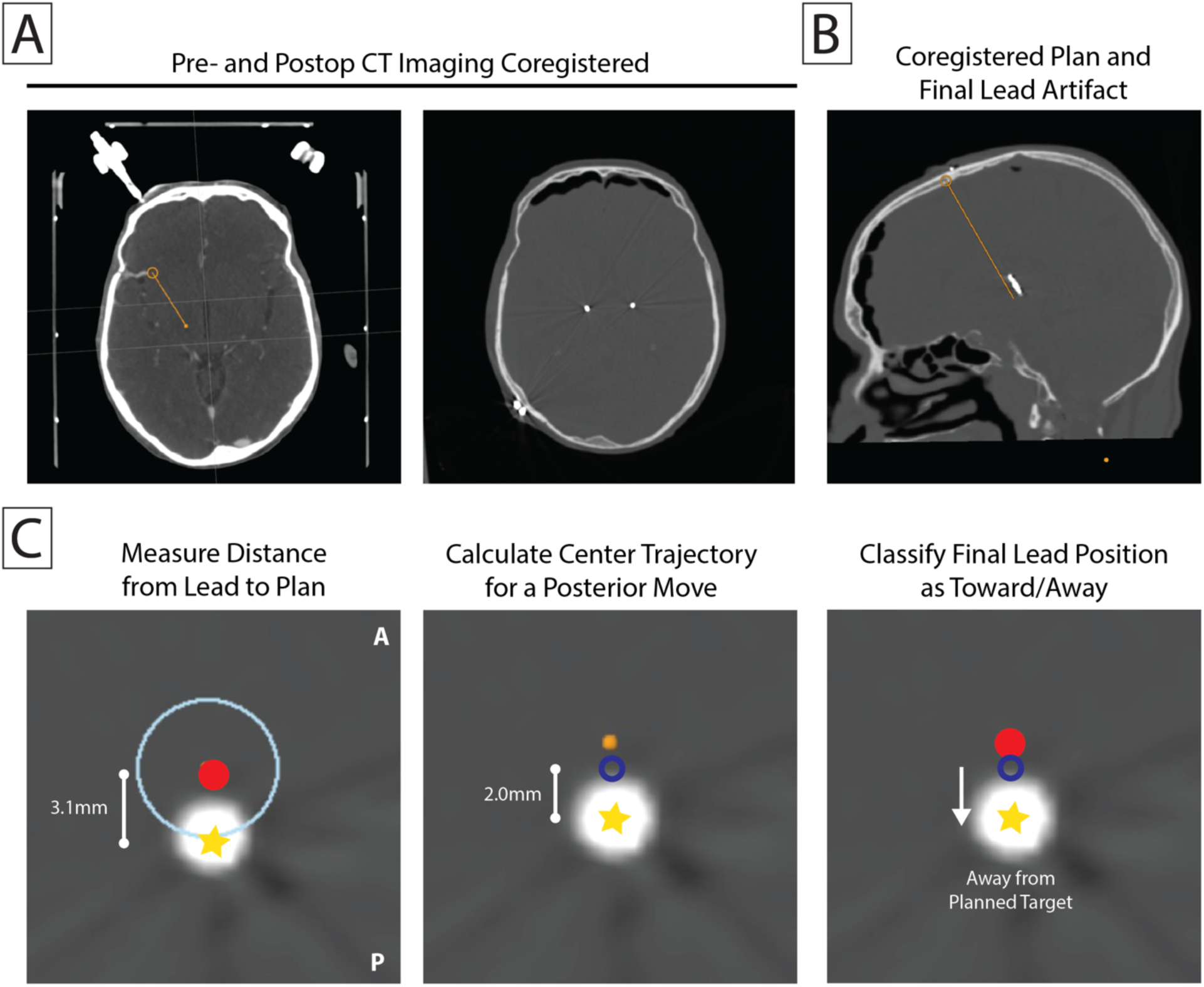
Co-registration workflow for clasficiation of physiology-guided trajectory adjustments. **(A)** Preoperative stereotactic CT used for surgical planning was rigidly co-registered with postoperative CT acquired on the day of surgery to localize the final DBS lead artifact. **(B)** The planned imaging-defined trajectory and the final implanted lead artifact are shown in axial and sagittal views following co-registration. **(C)** For cases implanted along a parallel trajectory, the distance from the final lead position (yellow star) to the imaging-defined target (red circle) was measured, and the position of the originally planned trajectory (calculated center trajectory; blue open circle) was reconstructed using the known 2-mm BenGun offset. Final lead placement was then classified as positioned closer to, farther from, or equidistant across the imaging-defined target. The example case shows a posterior move away from the planned target

### Data analysis

Surgical records were reviewed to categorize final lead placement as: (1) implantation at the planned depth along the originally explored trajectory; (2) implantation along the originally explored trajectory with a depth adjustment; or (3) implantation along a parallel trajectory. For parallel trajectory cases with suitable imaging, distances between the final lead position and the imaging-defined target were calculated, and the direction of deviation was classified as toward (suggesting correction of stereotactic error), away from, or equidistant across the target. All imaging analyses were performed independently of the operating surgeon and intraoperative neurologist (MRB).

For leads requiring trajectory adjustment, stimulation thresholds for near tremor resolution and persistent side effects were recorded both before (center track) and after trajectory adjustment when available. The therapeutic window was defined as the difference between the side-effect threshold and the near-resolution threshold. Descriptive statistics were calculated for post-adjustment stimulation parameters across all moved trajectories and reported as mean ± standard deviation. In cases with paired pre- and post-adjustment data, comparisons were performed using the Wilcoxon signed-rank test. Statistical significance was defined as p < 0.05.

## RESULTS

### Lead Adjustments

Among the 21 leads implanted along a parallel trajectory, 17 had preoperative trajectory data and postoperative imaging of sufficient quality for retrospective localization analysis. For these cases, the final lead position was compared with the imaging-defined target using co-registered imaging and reconstruction of the calculated center trajectory. Of the 17 analyzable parallel trajectory cases, 13 leads (76%) were repositioned further from the imaging-defined target than the calculated center trajectory, indicating divergence from the image-based plan. Two leads (12%) were repositioned an equal distance across the target, representing adjustments that also would not have been made based on imaging alone. Only two leads (12%) were repositioned closer to the imaging-defined target. The resulting final lead distances relative to the imaging-defined target ranged from 1.0 to 3.2 mm (Table 1, Fig. 4).

**Figure 4.**
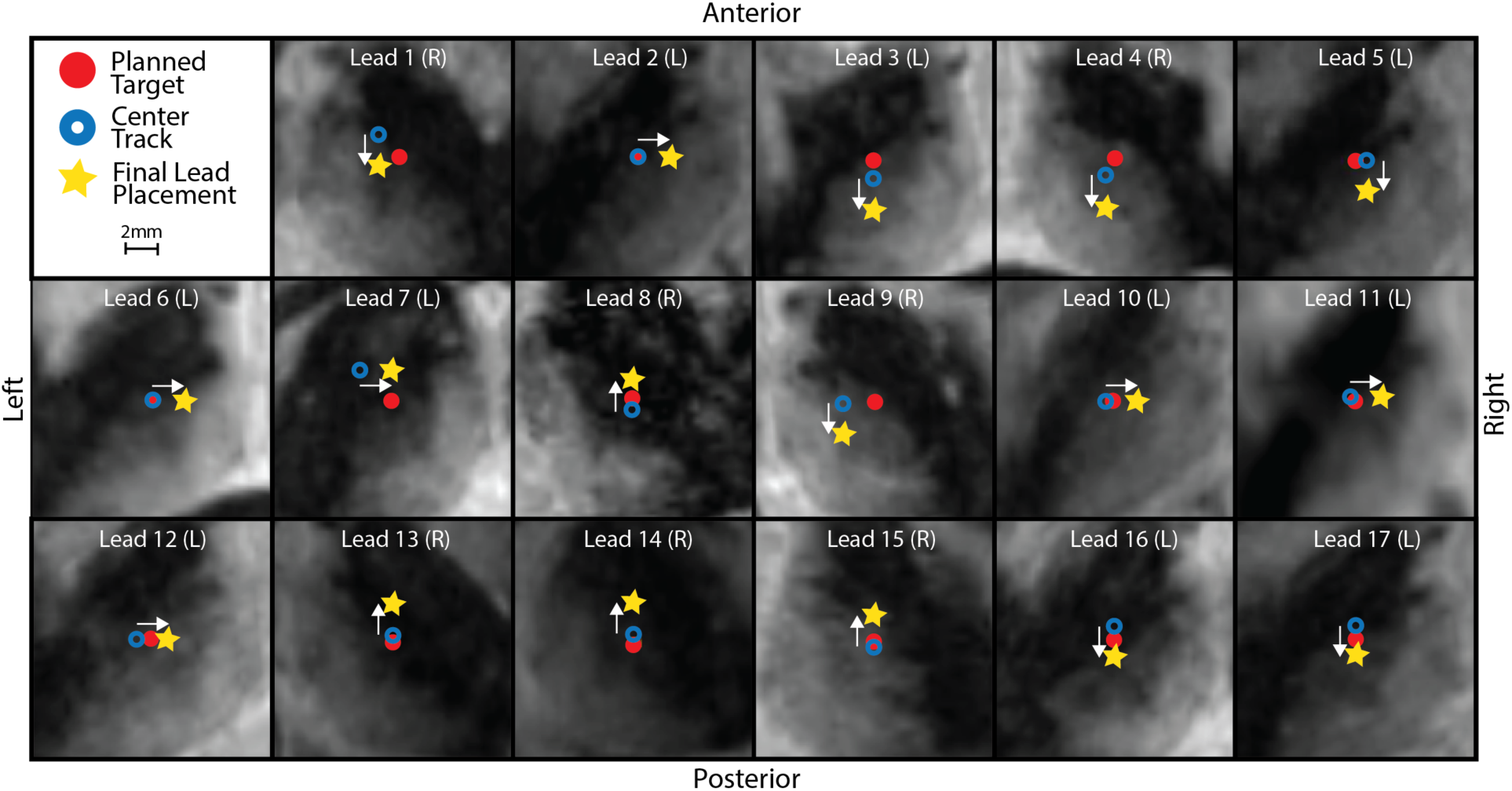
Direction and magnitude of physiologically-guided lead adjustments relative to planned target and calculated initial center track. White-matter nulled (FGATIR) sequence sliced orthogonal to the trajectory and at the midpoint of the stimulation electrode span for each of the 17 leads. These demonstrate the planned trajectory (red circle), the calculated center track (blue ring), and the final lead position after a move (yellow star). Arrows indicate the direction of the 2mm move in the BenGun. Only 2/17 leads were moved towards the intended target, while the remaining were either farther away (13) or equidistant (2).

Of the original 21 moved leads, the most common move direction was posterior (n = 8) and medial (n = 8), followed by anterior (n = 5); No lateral adjustments were observed. Improved single-unit recordings were cited as the primary reason for adjustment in 9 cases, including movement toward trajectories with tremor- or passive movement–tuned neurons (6/9) and away from trajectories with sensory-tuned activity (3/9). In 6 cases, intraoperative ring stimulation testing was the primary driver of adjustment, including movements away from stimulation-induced motor side effects consistent with internal capsule proximity (e.g., facial pulling) and movements toward trajectories with improved therapeutic window in the absence of sensory side effects. The remaining 6 leads were placed on non-center trajectories based on a combination of single-unit and stimulation findings. These directional patterns are consistent with expected anatomic–physiologic relationships: medial adjustments were directed away from the internal capsule, anterior adjustments away from the ventral caudal nucleus, and posterior adjustments were typically associated with improved therapeutic effect without eliciting persistent sensory side effects.

Among the 116 leads implanted along the planned trajectory, 67 required an adjustment in depth based on intraoperative physiologic findings. These depth adjustments included both superficial and deeper placements relative to the planned target depth with a mean displacement (± standard deviation) of 0.92 ± 0.49 mm.

### Macroelectrode Ring Stimulation Testing

Intraoperative ring stimulation data were available for 21 moved trajectories (Figure 5A), with paired pre- and post-adjustment testing in 12 cases. Across all moved trajectories, mean (± standard deviation) post-adjustment side-effect thresholds, near-tremor resolution thresholds, and therapeutic windows were 2.83 ± 0.55 mA, 0.99 ± 0.34 mA, and 1.85 ± 0.47 mA, respectively. In the subset with paired testing, trajectory adjustment was associated with significant improvement in stimulation characteristics. The amplitude required for near tremor resolution decreased following adjustment (Wilcoxon signed-rank, median = 2.00 vs 1.00; 95% CI = [-1.5 - 0.3]; p = 0.002), while the threshold for persistent side effects increased (median =1.82 vs 2.92; 95% CI = [0.5 2.0] p = 0.002). As a result, the therapeutic window significantly expanded following trajectory adjustment (median = 0.00 vs 1.77; 95% CI = [0.8 2.1]; p = 0.0005; Figure 5B). No leads in this cohort required revision following implantation.

**Figure 5.**
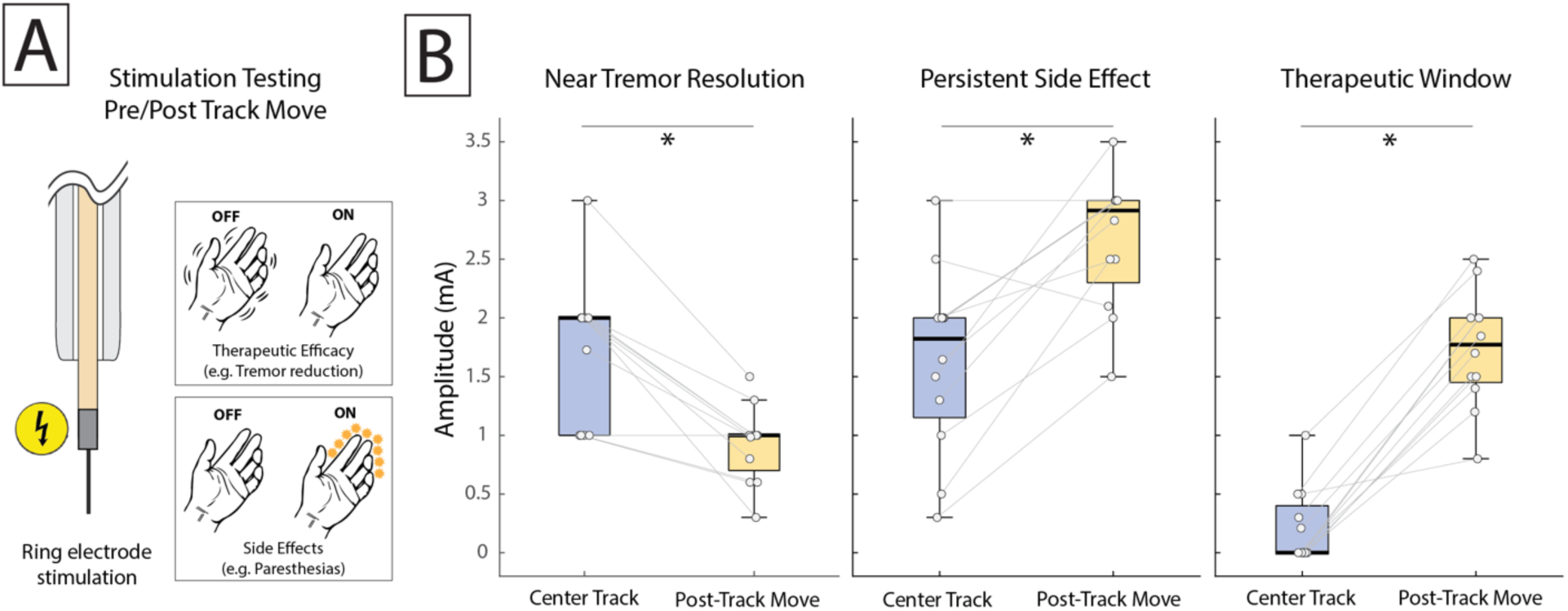
Intraoperative ring electrode stimulation testing before and after trajectory adjustment. **(A)** Schematic of intraoperative ring stimulation testing used to assess therapeutic efficacy (tremor reduction) and side effects. **(B)** Paired comparison of stimulation parameters before (center track) and after trajectory adjustment. Following adjustment, the amplitude required for near tremor resolution decreased, while the threshold for persistent side effects increased, resulting in a significant expansion of the therapeutic window (Wilcoxon signed-rank tests, all p < 0.05).

## DISCUSSION

In this retrospective series of awake Vim DBS procedures, we examined how often awake intraoperative physiologic testing prompted changes in lead placement and whether those changes converged with or diverged from the imaging-defined target. The principal finding is that parallel trajectory adjustments made according to intraoperative physiology and test stimulation more frequently resulted in final lead positions farther or equidistant from, rather than closer to, the planned imaging target. The vast majority (88%) of off-center moves would not have been made had we relied on intraoperative imaging feedback alone. This observation suggests that imaging-based planning and intraoperative physiologic guidance may not always identify the same optimal stimulation site at the individual patient level.

We argue that awake DBS with MER remains a practical and valuable approach in the era of advanced imaging. While modern asleep workflows leverage high-resolution MRI, tractography, and intraoperative volumetric imaging for targeting^4,7^, these tools can also be used in awake workflows and do not replace the unique information gained from real-time awake physiologic feedback. Importantly, the use of an awake protocol should not be conflated with cutting corners elsewhere: all patients still undergo the full battery of preoperative imaging, and we maintain rigorous intraoperative imaging standards, using O-arm volumetric imaging before closure in asleep cases, and fluoroscopy with immediate postoperative CT in awake cases. This combined approach underscores that awake testing is not a legacy technique competing with modern imaging, but a complementary method that refines placement based on patient-specific physiology in ways that imaging alone cannot.

### Value of real-time clinical and physiologic feedback

A central feature of awake DBS surgery is the ability to directly observe stimulation-induced therapeutic effects and side effects at the time of implantation allowing the surgical team to balance benefit and adverse effects during lead placement, a process that cannot be replicated using imaging outcomes alone^15^. Even if microlesional effects from lead insertion confound assessment of efficacy, stimulation-induced side effects often remain^16^. In awake workflows, repositioning decisions are therefore informed by direct clinical and physiologic observation, whereas in asleep workflows such judgments rely on inferred stimulation effects at an imaging-defined anatomic location.

In the present series, only 2 of the 17 parallel trajectory adjustments would have been prompted by intraoperative imaging findings. The remaining adjustments reflected responses to intraoperative physiologic and clinical feedback, illustrating how awake testing can influence lead placement in ways not predicted by preoperative imaging. Depth optimization was also common. Contemporary segmented DBS contacts span a limited axial distance (approximately 3.5 mm), such that small differences in depth can determine whether directional steering can be effectively leveraged near the physiologic target^17,18^. Imaging alone cannot determine whether the most responsive physiologic zone within a target is centered within the restricted span of the contact array. Awake MER provides an opportunity to make these fine depth adjustments in real time. Notably, there are newer generation electrodes with larger directional spans that may further improve depth optimization.

Most patients experience meaningful clinical benefit from DBS regardless of whether surgery is performed awake or asleep^19–21^, and the present study does not seek to compare outcomes between surgical paradigms. Instead, these findings highlight how intraoperative physiology can influence lead placement decisions in a subset of cases when imaging-defined targets and functional responses are not fully concordant. As DBS technologies continue to evolve toward greater spatial precision and adaptive stimulation strategies, intraoperative physiologic recordings may play a complementary role alongside imaging by helping localize functionally relevant regions or verify the presence of patient-specific biomarkers at the time of implantation^22–24^.

### Why might an ‘optimal’ imaging outcome not produce optimal therapy?

Even with meticulous planning and high-resolution imaging, the functionally optimal site for DBS may not align precisely with the planned anatomical coordinates. First, there is natural variability in the spatial representation of somatotopy and tracts within nuclei^25,26^. Second, neuronal elements associated with stimulation-induced therapeutic effects may interdigitate with those associated with side-effect, in which case side effects may emerge even at seemingly ideal anatomic targets^27^. Third, structural variation across individuals, particularly at borders like the Vop–Vim transition, is not reliably visible on current imaging^28^. While this limitation is especially pronounced in Vim, similar (though likely less prominent) discordance between imaging- and functionally-defined targets may also occur in other DBS targets, where functional subregions and laminar structures may not be fully captured on routine imaging. Finally, the brain itself deforms in response to the advancing rigid cannula, and the surrounding tissue strains and displaces around it, so even a perfectly aligned trajectory can deviate subtly from the intended anatomy which may or may not be appreciated on intraoperative imaging. These realities suggest that MER-guided adjustments are deliberate refinements based on functional data that imaging cannot provide.

### When awake DBS may not be appropriate

Awake DBS with MER requires active patient engagement for optimum benefits. Excessive anxiety, including claustrophobia, can make the experience intolerable and undermine cooperation during testing^21,29^. Cognitive impairment can similarly limit cooperation. Severe axial dystonia, tics, or fixed painful postures can increase the chance of frame slippage or injury with awake surgery. Some targets do not offer immediate behavioral feedback and lie far from structures producing acute side effects, such as the anterior nucleus of the thalamus (ANT) for epilepsy and therefore cannot benefit from awake testing. However, even in cases were asleep implantation is preferred, MER can still provide useful information regarding structural borders.

### Limitations

This retrospective, single-practice study may have limited generalizability. Reconstruction of the center trajectory is inferred and relies on stereotactic geometry and co-registered imaging, and cannot fully account for subtle intraoperative brain deformation or possible deviations between the microelectrode and permanent lead^30^. Analyses were restricted to thalamic targets in which the initial rigid cannula was left in place during parallel trajectory testing, helping preserve the stereotactic framework during track moves. Delayed postoperative imaging may reduce some of these sources of uncertainty, although any residual error would be expected to introduce small spatial variability rather than a systemic directional bias.

## CONCLUSION

The present findings provide insight into contemporary DBS practice by quantifying how often intraoperative physiologic testing influences lead placement in ways not predicted by imaging alone. In a subset of cases, awake physiology and testing resulted in lead adjustments that would not have been indicated by imaging alone, and improved patient therapeutic windows, underscoring that imaging and physiology can provide non-identical but complementary information during DBS surgery. Understanding when such discordance arises may inform surgical planning and intraoperative workflows as DBS technologies continue to evolve toward greater spatial precision and individualized stimulation strategies.

## Data Availability

All data produced in the present work are contained in the manuscript.

## Author Contributions

MRB, BTK, and KJM conceived the study, collected the data, and edited the manuscript. MRB and BTK performed data analysis, created figures, and wrote initial manuscript drafts. KJM secured funding.

## Abbreviations

DBS: Deep brain stimulation
MER: Microelectrode recordings
Vim: Ventral intermediate nucleus

